# Outcome of Graves’ Disease and Associated Genetic Predisposition: A Systematic Review of Literature

**DOI:** 10.1101/2024.09.27.24314519

**Authors:** Md. Ikhsan Mokoagow, Ahmad Zufar Manthovani, Marina Epriliawati

## Abstract

**Background:** Graves’ disease (GD) is an autoimmune hyperthyroidism that primarily affects the thyroid gland. The susceptibility, activity, and risk of recurrence of GD may be influenced by various genetic factors, which can interact with environmental triggers to precipitate disease relapse. This review examines the literature on the genetic predisposition to GD recurrence.

**Methods:** Literature from PubMed, EBSCOhost, and SCOPUS were searched. Three reviewers performed study selection and critical appraisal.

**Result:** Five studies were selected. The literature indicates a risk of relapse in PTPN22, CTLA-4, TSH-R, IL-21, KREC, and HLA gene abnormalities.

**Conclusion:** Several genes are associated with GD relapse. However, predicting relapse risk requires consideration of multiple factors beyond genetic predisposition.

## 1. Introduction

Graves’ disease (GD) is the most common cause of hyperthyroidism, which primarily affects the thyroid gland. It is an autoimmune disease that is precipitated by environmental factors like exposure to iodine, stress, smoking, infection, postpartum, and also immune reconstitution after *Highly Active Antiretroviral Therapy* (HAART). Graves’ disease accounts for 20-30 annual cases per 100,000 individuals in iodine-rich parts of the world every year, which affects 3% of women and 0.5% of males all over the world.^1,2^

The Predisposition to GD appears to be influenced by multiple genes. It is considered as Human Leukocyte Antigens (*HLA*) *class II*-associated disorder, and several genes are also involved in the development of Graves’ disease such as *CTLA-4, TSHR*, Thyroglobulin gene (*Tg*), *CD40, PTPN22, CD25, FOXP3*, and *VDR*.^3–8^ GD can be managed through antithyroid drugs (ATD), radioactive iodine therapy or surgical intervention. While antithyroid drugs effectively control the symptoms of thyrotoxicosis, they do not address the underlying cause of GD, leading to frequent relapses.^1,9^

In individuals with a genetic predisposition, specific environmental risk factors exposure can trigger the production of autoantigens against the Thyroid Stimulating Hormone (TSH) receptor, leading to the development or relapse of the disease.^10^ We aimed to systematically review the available research on the genetic predisposition to relapse of GD, providing a brief overview of the genes involved in disease relapse, its mechanisms, and prediction of relapse.

## 2. Method

Initially, we searched for reviews on the outcome related to the relapse of Graves’ disease and its associated genetic predisposition on the International Prospective Register of Systematic Reviews (PROSPERO). We found no existing systematic review. As the Preferred Reporting Items for Systematic Review and Meta-Analysis (PRISMA) statement recommended, this systematic review was registered on PROSPERO (www.crd.york.ac.uk/PROSPERO).

We conducted a literature search on April 4th, 2024, using PubMed, EBSCOhost, and Scopus with the keywords from Table 1. The search was restricted to English published within the last ten years. We aimed to identify all observational, comparative, meta-analyses, and systematic reviews. A total of 12 studies were identified. Three reviewers carried out the study selection and critical appraisal to assess eligibility. Seven studies were excluded due to having different objectives. Hence, five studies were included. The selected articles were critically reviewed using the Newcastle-Ottawa Scale (NoS). The corresponding flowchart is shown in Figure 1.

**Table 1.**
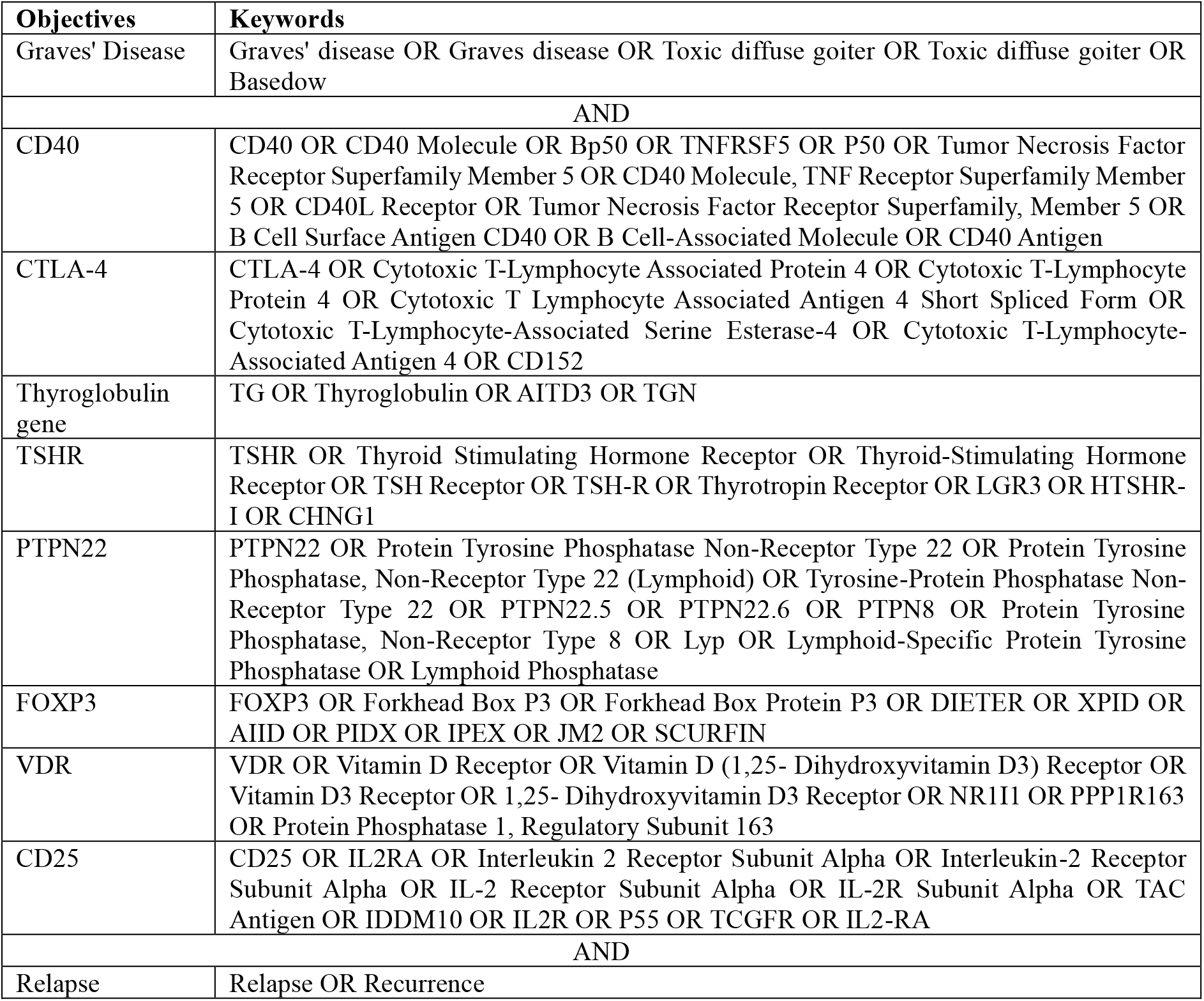
Keywords for advance searching.

**Figure 1.**
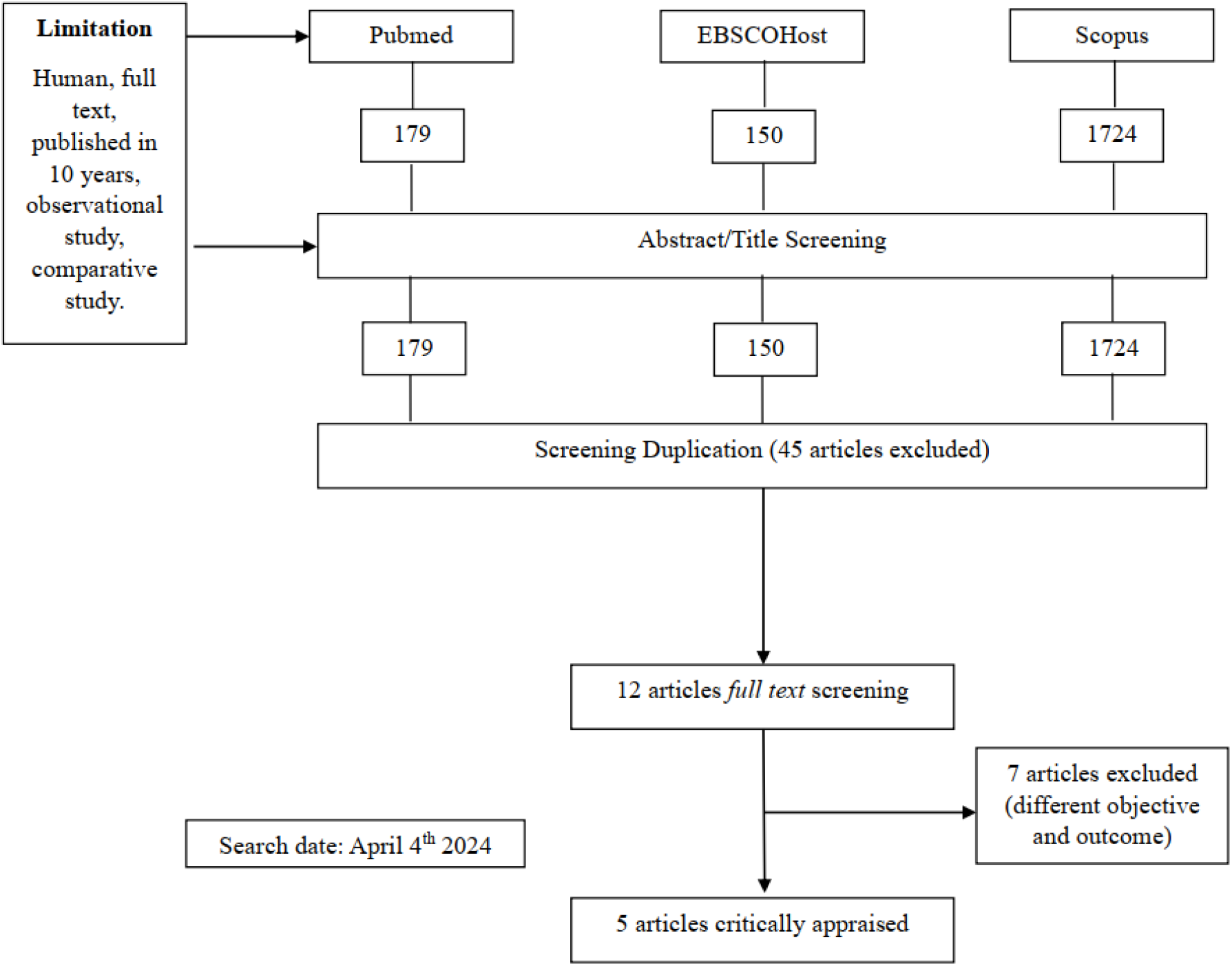
Study’s method flowchart

## 3. Result

Five studies were selected, comprising 2 retrospective cohort studies, 1 prospective cohort study, and 2 case-control studies. The selected articles showed a risk of recurrence in *PTPN22, CTLA-4, TSH-R, IL-21, KREC*, and *HLA* gene abnormalities (Table 2). However, no studies within the specified period supported an association with recurrence for other genes. Literature assessment using the Newcastle-Ottawa Scale, converted to the Agency for Healthcare Research and Quality (AHRQ) standards, indicated that all selected studies were of good quality (Table 3).

**Table 2.**
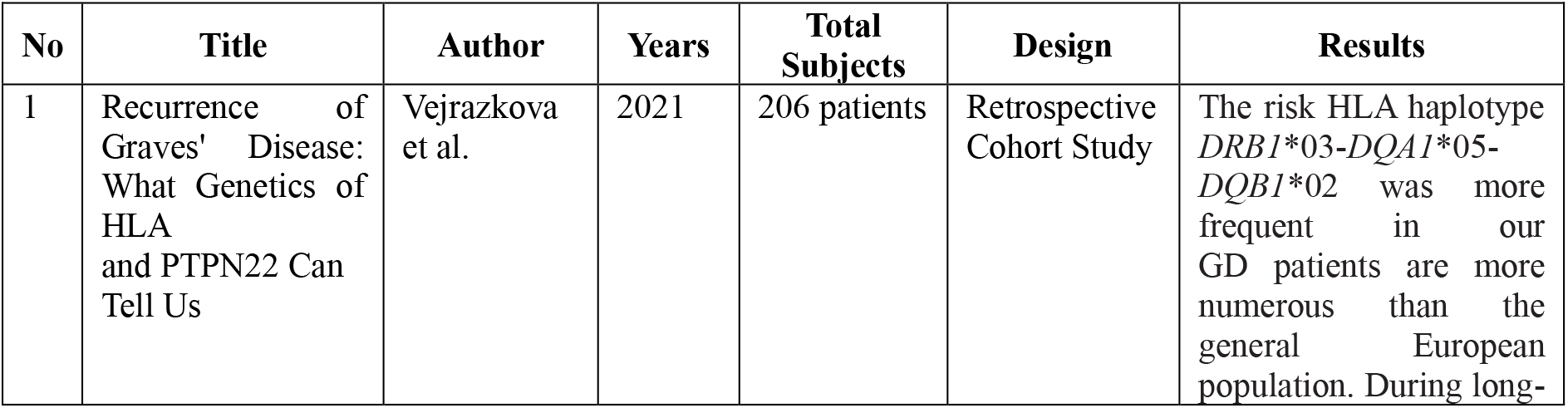
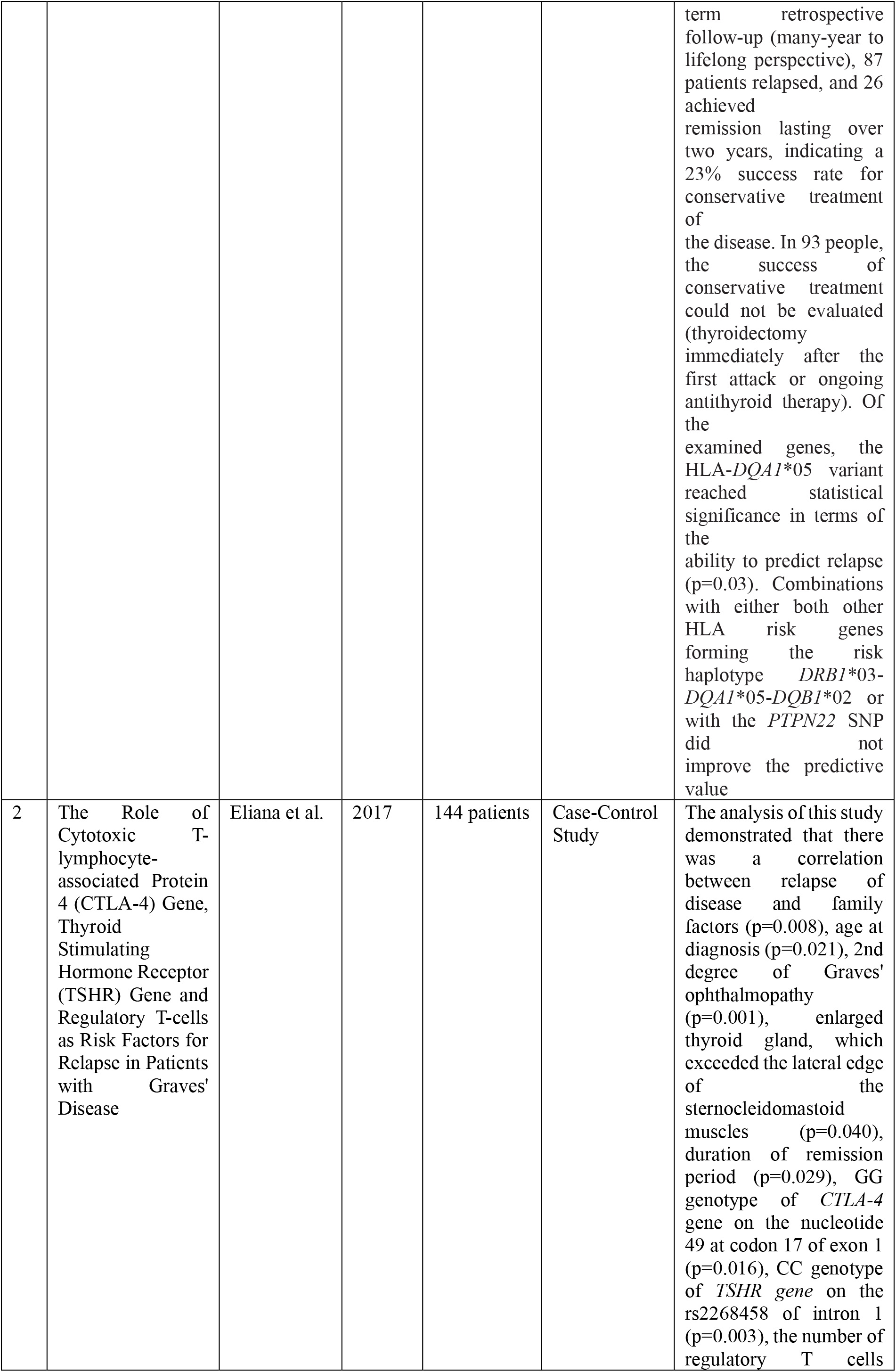
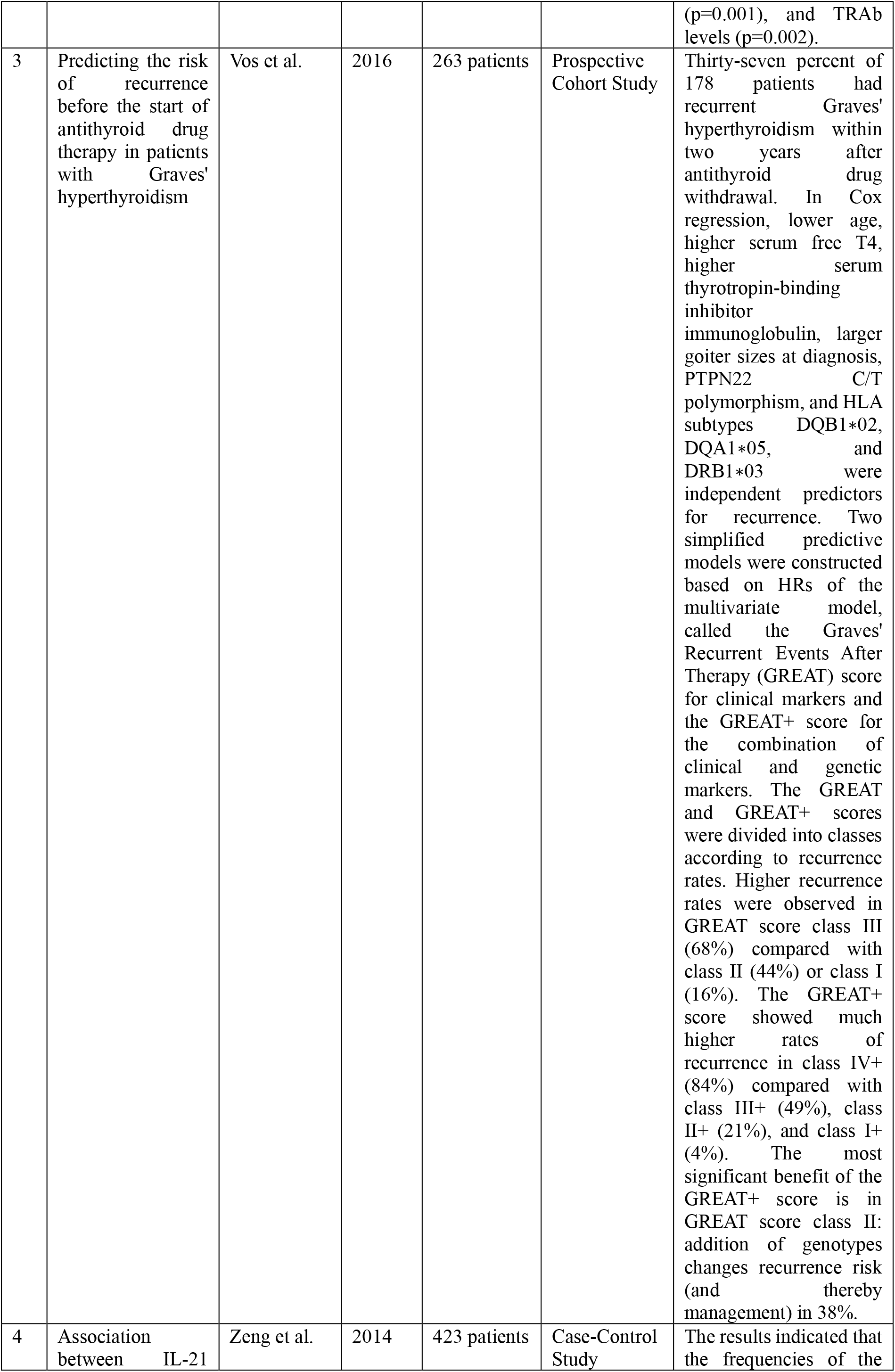
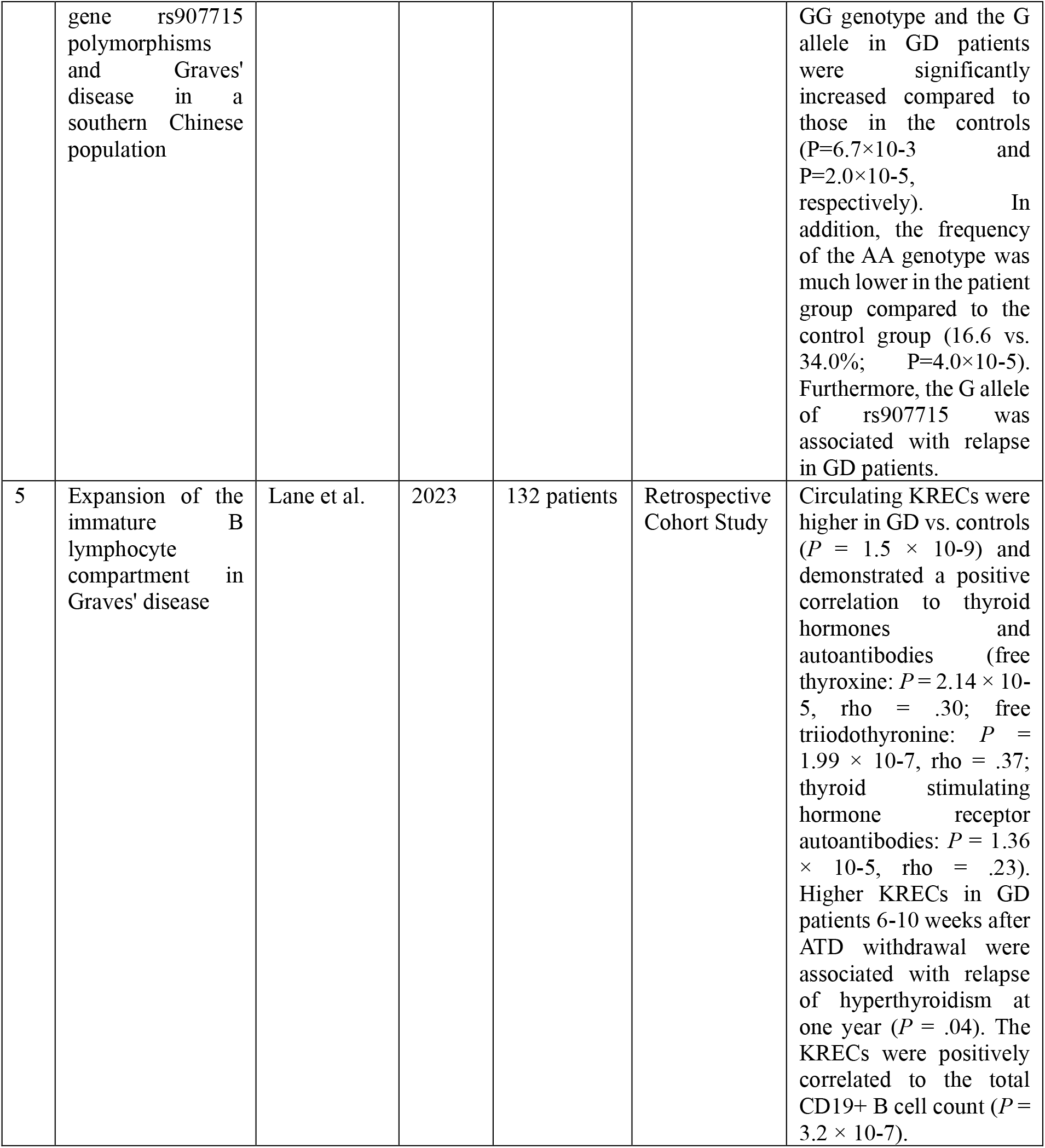
Selected-studies description.

**Table 3.**
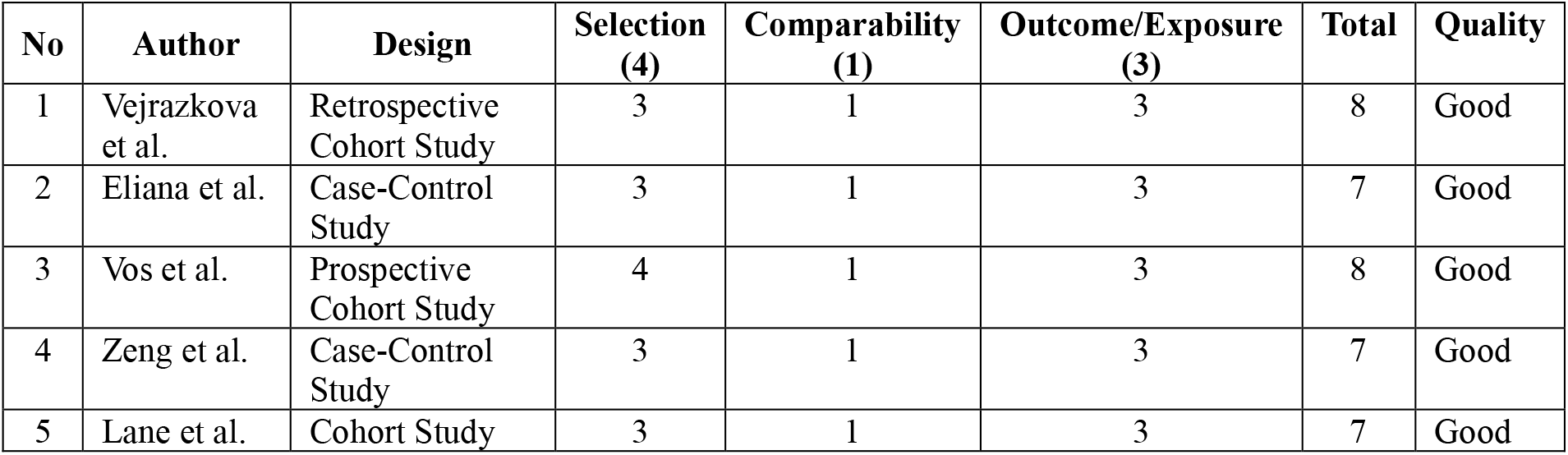
New Castle Ottawa Scape and Quality Assessment.

## 4. Discussions

### 4.1. Graves’ Disease Relapse

Relapse in GD is defined as the reappearance of hyperthyroidism symptoms, along with laboratory results showing serum T_4_ and/or T_3_ levels above the normal range. Relapse occurs most likely within the 6-12 months or later after the withdrawal of ATD. All patients should be monitored for relapse during the first year after completing treatment and then annually thereafter. Patients with severe hyperthyroidism, large goiters, or consistently high TSH-R-Antibody titers are at the highest risk of relapse after treatment cessation.^2,11^

### 4.2. Human Leukocyte Antigen (HLA) Complex

The Human Leukocyte Antigen (*HLA*) complex is associated with the autoimmune nature of GD. The *HLA Class II region*, located within the Major Histocompatibility Complex (MHC) on chromosome 6p21, plays a crucial role in the autoimmune process. *HLA* class II molecules are expressed on the surface of B cells, macrophages, dendritic cells, and activated T cells. They are involved in the processing and presentation of exogenous antigens. This region is highly polymorphic and considered critical in developing autoimmune diseases. Within the *HLA*, the DR3 haplotype (*DQB1*02, DQA1*0501, DRB1*03*) predisposes to GD due to strong linkage disequilibrium among the *DQA1, DQB1*, and *DRB1* loci. In contrast, the DR7 haplotype (*DQA1*0201, DQB1*0302, DRB1*07 or DQA1*0201, DQB1*02, DRB1*07*) appears to offer protection (Antonelli et al). ^3,12,13^

Vos et al., in the prospective cohort study, stated that HLA Class II haplotypes DRB103, DQA105, and DQB102 are well-established as being associated with an elevated risk of developing Graves’ hyperthyroidism in Caucasians. The research has identified that these specific polymorphisms—*HLA DRB103, DQA105*, and *DQB102*—are significant predictors of recurrence following antithyroid drug therapy. Another study has examined the association between recurrence risk and *HLA DQA1*05* and found no significant correlation. (Vos et al.).^12^

The *DRB1*03, DQA1*05*, and *DQB1*02* allelic groups groups have proven to be predictors of the development and recurrence of GD. The *HLA-DQA105* variant and the *DRB103-DQA105-DQB102* haplotype were more common in GD patients than in the general European population. However, the gene tested individually is not a reliable predictor of relapse in the Czech population. A comparison with some reference European populations suggests a higher minor allele frequency in the GD patients. The *HLA-DQA1*05* allelic group was significantly associated with relapse or prolonged unsuccessful conservative treatment (Vejrazkova et al.).^10^

### 4.3. PTPN22

The Protein Tyrosine Phosphatase-22 (*PTPN22*) gene encodes the lymphoid tyrosine phosphatase (*LYP*) protein, which acts as a potent negative regulator of T-cell receptor signaling. *PTPN22* plays a role in the pathogenesis of GD. The *PTPN* C/T Single Nucleotide Polymorphism (SNP) (*rs2476601*) is also associated with Graves’ hyperthyroidism and recurrence rate (Vos et al). The *PTPN22* genetic variant *rs2466601* has been identified as a potential predictor of Graves’ disease and was included in the Graves’ Events After Therapy (GREAT+) score and *HLA* variants. However, the study by Vejrazkova et al. did not demonstrate the predictive potential of the *PTPN22* variant for disease recurrence. Therefore, this gene cannot be relied upon to predict relapse in the Czech population.^10,12–14^

The *PTPN22 R620W* SNP is associated with Graves’ disease and causes a functional change in the *LYP* protein. The tryptophan-bearing LYP allele cannot bind with C-terminal src kinase (Csk), an inhibitory complex partner that regulates key T cell receptor signaling kinases. Paradoxically, this mutation enhances the protein’s ability to inhibit T cells, making it a gain-of-function mutation. This reduced T cell signaling may allow self-reactive T cells to escape thymic deletion and persist in the periphery. Ethnic studies have shown varying associations, likely due to founder effects or the absence of specific variants in different populations. For instance, the tryptophan variant of the *PTPN22* gene is not present in the Japanese population, suggesting it does not contribute to autoimmunity in this group (Jacobson et al.).^5^

### 4.4. CTLA-4

Cytotoxic T Lymphocyte-Associated Molecule 4 (*CTLA-4*) delivers a negative signal to T cells, regulating their transition to a state of non-responsiveness and tolerance, thereby controlling their proliferation. Polymorphism in the gene promoter, which controls gene transcription, may affect the *CTLA-4* level by changing the binding of transcription factors.^15,16^

The *CTLA-4* gene consists of 3 introns and four exons. The Exon 1 of *CTLA-4* is demonstrated to have a role as a risk factor and relapse of autoimmune disease. The GG genotype of *CTLA-4* at nucleotide 49, codon 17 of exon one, is known to have a higher risk of relapse (Eliana et al.). The study from Wang et al. confirms that the *CTLA-4* genotypes are useful in predicting the relapse of GD after treatment cessation.^15,16^

### 4.5. TSHR

The *TSHR* gene has long been considered a prime candidate gene for Graves’ disease due to the presence of stimulating TSH receptor (*TSH-R*) autoantibodies, a hallmark of the disease. The *TSH-R* is expressed on the surface of thyroid epithelial cells and is located on the long arm of chromosome 14 (14q31). It binds to Thyroid Stimulating Hormone and triggers the production of thyroid hormones. Intron 1 of the *TSHR* gene functions as a regulator of RNA, influencing the initiation site for mRNA formation. Research findings have been inconsistent regarding the association between the *TSHR* gene and GD (Jacobson et al). However, a study by Eliana et al. found that GD patients with the CC genotype for the *TSHR* gene SNP *rs2268458* in intron 1 had a higher risk of relapse compared to those with the TT genotype.^5,15^

### 4.6. IL-21

Interleukin-21 (*IL-21*) is located on chromosomes 4q26-27 and 16p11, close to *IL-2*, a region associated with autoimmune thyroid diseases. *IL-21* has been shown to play a crucial role in interactions among B cells, T cells, NK cells, and dendritic cells, potentially serving as a key cytokine linking innate and adaptive immune responses. Zhang et al. found a significant association between Graves’ disease (GD) and polymorphisms in the *IL-21* gene region. Zeng et al. specifically identified that the *rs907715* variant in intron 3 of the *IL-21* gene is linked to GD susceptibility. Individuals carrying risk-associated G alleles are more susceptible to GD and tend to relapse after treatment cessation. The biological mechanisms underlying the G allele’s effects remain poorly understood.^17–19^

### 4.7. KREC

Kappa-deleting recombination excision circles (*KRECs*) are formed during B-cell development in cases where B lymphocytes fail to successfully rearrange the Ig kappa light chain genes (*IGK*) after the Ig heavy chain rearrangement. This results in the deletion of the constant kappa gene segment (*IGKC*) through recombination with the kappa-deleting element (*Kde*), a sequence located about 24 kb downstream. The recombination deletes portions of DNA, and the excised ends form circular DNA elements known as *KRECs*. Normally, cells with receptors for autoantigens and their *TRECs* or *KRECs* are deleted in central lymphoid organs after interacting with local epithelial cells. If central immune tolerance is impaired, many autoreactive T and B cells, along with their *TRECs* and *KRECs*, escape into circulation and target various organs, increasing the risk of autoimmunity.^20,21^

*KREC* levels have been linked to disease status and activity in another B-cell-mediated autoimmune disorder. This suggests that *KRECs* might offer mechanistic insights into the immunopathology driving autoimmunity in Graves’ disease and could also be associated with relevant clinical phenotypes or outcomes. A study conducted by Lane et al. is the first to report that higher *KREC* levels are associated with GD relapse or disease activity.^22^

### 4.8. Other genes related to relapse of Graves’ Disease

Other genes are found related to relapse, such as *CD40* with 3 polymorphisms (*rs745307, rs11569309*, and *rs3765457*). *CD40*, a crucial co-stimulatory molecule on antigen-presenting cells, plays a major role in T cell-dependent B cell activation. Polymorphisms that increase *CD40* expression could potentially trigger GD relapse by boosting the humoral response. Haplotype studies link *CD40* polymorphisms associated with GD outcomes to higher *CD40* mRNA expression levels.^23^

The *TG* gene encodes thyroglobulin, a protein essential for thyroid hormone production. Elevated serum thyroglobulin levels are often observed in GD and have been linked to a higher relapse risk. Genetic variants in thyroid-specific genes may interact with immune-related genes, potentially influencing GD outcomes synergistically. *CD25, VDR*, and *FOXP3* genes are associated with the susceptibility to developing Graves’ Disease. However, there is no specific information about these genes being associated with relapse of GD.^4,6,8,23^

In conclusion, genes associated with Graves’ disease relapse include *HLA, PTPN22, CTLA-4, TSHR, IL-21*, and *KREC*. While detected mutations in these genes may increase the risk of recurrence, genetic testing alone cannot be solely relied upon to predict relapse in an individual. Multiple factors need to be considered when predicting Graves’ disease recurrence. Additionally, the tendency for each gene mutation varies based on geographical distribution, with the frequency and form of mutations differing across countries. In this review, we found limited recent research within the past ten years, which restricted the literature included in this systematic review. Further research on the genetic predisposition to Graves’ disease relapse is necessary so that, in the future, genetic mapping in Graves’ disease patients can more accurately predict outcomes.

## Data Availability

All data produced are available online at MedLine, EBSCOhost and Scopus

